# Electrocardiogram sleep staging on par with expert polysomnography

**DOI:** 10.1101/2023.10.13.23297018

**Authors:** Adam M. Jones, Laurent Itti, Bhavin R. Sheth

**Affiliations:** Neuroscience Graduate Program, University of Southern California, Los Angeles, California; Department of Electrical & Computer Engineering, University of Houston, Houston, Texas

**Keywords:** sleep, stages, polysomnography, electrocardiogram, cardiosomnography, deep learning

## Abstract

Accurate classification of sleep stages is crucial in sleep medicine and neuroscience research, providing valuable insights for diagnoses and understanding of brain states. The current gold standard for this task is polysomnography (PSG), an expensive and cumbersome process involving numerous electrodes, often performed in an unfamiliar clinic and professionally annotated. Although commercial devices like smartwatches track sleep, their performance compares poorly with PSG. To address this, we present a neural network that achieves gold-standard levels of agreement using a single lead of electrocardiogram (ECG) data (five-stage Cohen’s kappa = 0.725 on subjects 5 to 90 years old). Our method offers an inexpensive, automated, and convenient alternative. Cardiosomnography, or a sleep study conducted with electrocardiography only, could take expert-level sleep studies outside the confines of clinics and laboratories and into realistic settings. This would render higher-quality studies accessible to a broader community, enabling improved sleep research and sleep-related healthcare interventions.

## Introduction

The classification of sleep stages aids in the understanding of brain states and unconscious processes^1^. Stages, as formalized in 1968 by Rechtschaffen and Kales (R&K)^2^, and later revised in 2007 by the American Academy of Sleep Medicine (AASM)^3^, are divided into Wake, rapid eye movement (REM), and Non-REM (NREM) stages 1 through 3 (N1, N2, N3). These stages occur in a cyclic pattern with specific EEG signatures and correlate with physiological and neurological processes, such as waste clearing and particular types of memory consolidation^4,5^. Currently, clinically relevant sleep staging, or sleep stage scoring, requires polysomnography (PSG). During PSG, a patient or subject typically spends one or more nights in a clinic during which they wear electrodes that collect at least a dozen channels of data, including brain activity (electroencephalography, EEG), eye movement (electrooculography, EOG), muscle tension (electromyography, EMG), ECG, and respiration. Afterward, an experienced paid human scorer annotates the night to assess the stages and other pertinent notations. Because it can be expensive—taking approximately two hours to annotate a single night of sleep—automated methods are increasingly used^6,7^.

With the proliferation of commercial health-related sensors, spurred on by the public’s interest in self-tracking, commercial sleep trackers have exploded in popularity^8^. Moreover, the increasing understanding of the importance of sleep has fueled the healthcare community’s interest in sleep monitoring for precision medicine^9^. The cost of PSG (human labor and equipment) and sheer cumbersomeness for the sleeper (wearing electrodes, while non-invasive, is intrusive and inconvenient, leading to unrepresentative data) make it a non-starter for widespread adoption in the laboratory and beyond the laboratory setting.

Therefore, to fill this void, methods that measure other aspects of bodily function, such as ECG, respiration, heart rate variability, etc., have sprung up in recent years. Two advantages some of these methods have—specifically ones that use ECG—over EEG is that they require fewer electrodes (three versus dozens), and ECG is a much stronger signal than EEG. Whereas the performance of these commercial devices is promising, there is significant room for improvement; they often ignore harder-to-classify stages, exclude subjects based on age or health, and agree poorly with human-scored PSG—the “gold standard”. The ideal “sweet spot” outcome is a cheap, patient-friendly method that agrees with classical PSG; this would open the door to more research and clinical applications.

Broadly speaking, two issues have hampered efforts to score sleep stages automatically at an expert level without the full complement of biophysical inputs that PSG requires. First, researchers originally defined sleep stages by their manifestations in EEG, EOG, and EMG. The assumption that these inputs are, by definition, necessary has impeded the search for equally informative surrogates. Moreover, until recently, the second obstacle was the lack of enormous datasets for training generalizable classifiers. Today, tens of thousands of recordings of expert human-scored PSG are freely available, which makes it possible to overcome both issues. We investigated ECG specifically because it is a much more robust signal than EEG and because many PSG datasets collect it, albeit often as an afterthought. **Specifically, we set out to determine if it is possible to score sleep stages as well as human-scored PSG using only a single lead of ECG data.**

Here we demonstrate that by using a neural network trained on a large sleep dataset, it is possible to achieve gold-standard levels of agreement with PSG using only ECG data. We developed a neural network that uses a single lead of ECG data as the only biophysical input to score sleep stages, i.e., using none of the main inputs in classical PSG (e.g., EEG, EOG, and EMG). To improve the generalizability of the resulting model, we trained on a dataset of several thousand recordings from subjects 5 to 90 years old to approximate the U.S. census.

Furthermore, we developed a new loss function to align the training objective with the standard performance metric for comparing sleep scoring, namely Cohen’s kappa. Finally, we show that our method is a significant improvement over current commercial devices for sleep staging.

## Results

We present our model and results in four sections. First, we introduce the source datasets, processing pipeline, and deep neural network. Next, we compare our network’s performance against expert human-scored PSG and other EEG-less models (i.e., sleep staging models that exclude EEG as an input) and show results from analyses of the testing set. Then we analyze the impact on the network’s performance of various modifications. Finally, we demonstrate the real-time performance of such a network. The approach and results below support the claim that ECG is suitable for the highest-quality sleep studies and introduces a network that could make cardiosomnography-based sleep studies widely available.

### Scoring sleep with ECG

We demonstrate the broad applicability of our network (Fig. 1a) by scoring sleep stages from recordings taken from five different studies (Fig. 1b)—chosen for their substantial age range (5-90yr. = decades 1-10). We processed each recording from the original studies to calculate various quality measures (e.g., signal source, missing data, artifacts, etc.) about the provided data and to harmonize the data (e.g., standardize the sampling rate and normalize the amplitudes). We discarded recordings that did not meet the quality metrics defined in Methods. Our pipeline then randomly selected four thousand recordings (3,000 testing, 500 validation, and 500 testing) such that (given the limitations in the datasets), the distribution of subjects matched the 2022 U.S. census estimates across decades as well as the mean age (Fig. 1f). Additionally, we mirrored the overall distribution of age, sex, and recording source, across the three sets (i.e., training, validation, and testing). As desired by selecting a broad range of subjects, the recordings contained a wide distribution in stage ratios (i.e., the percent of the night spent in a specific stage), including a substantial number of recordings that had no epochs containing N1, N3, or both (Fig. 1c). Additionally, there is substantial variability in recording lengths, from 5.5 to 14.3 hours (Fig. 1d). When stratifying the recordings by decade, there are noticeable age-dependent shifts in the stage ratios (Fig. 1e). Both the likelihood of time-dependent changes in stage ratios (e.g., N3 is predominantly seen early in the night, while REM is predominantly seen at the end) and the age-dependent shifts (e.g., N3 decreases with age) were expected^10,11^.

**Figure 1.**
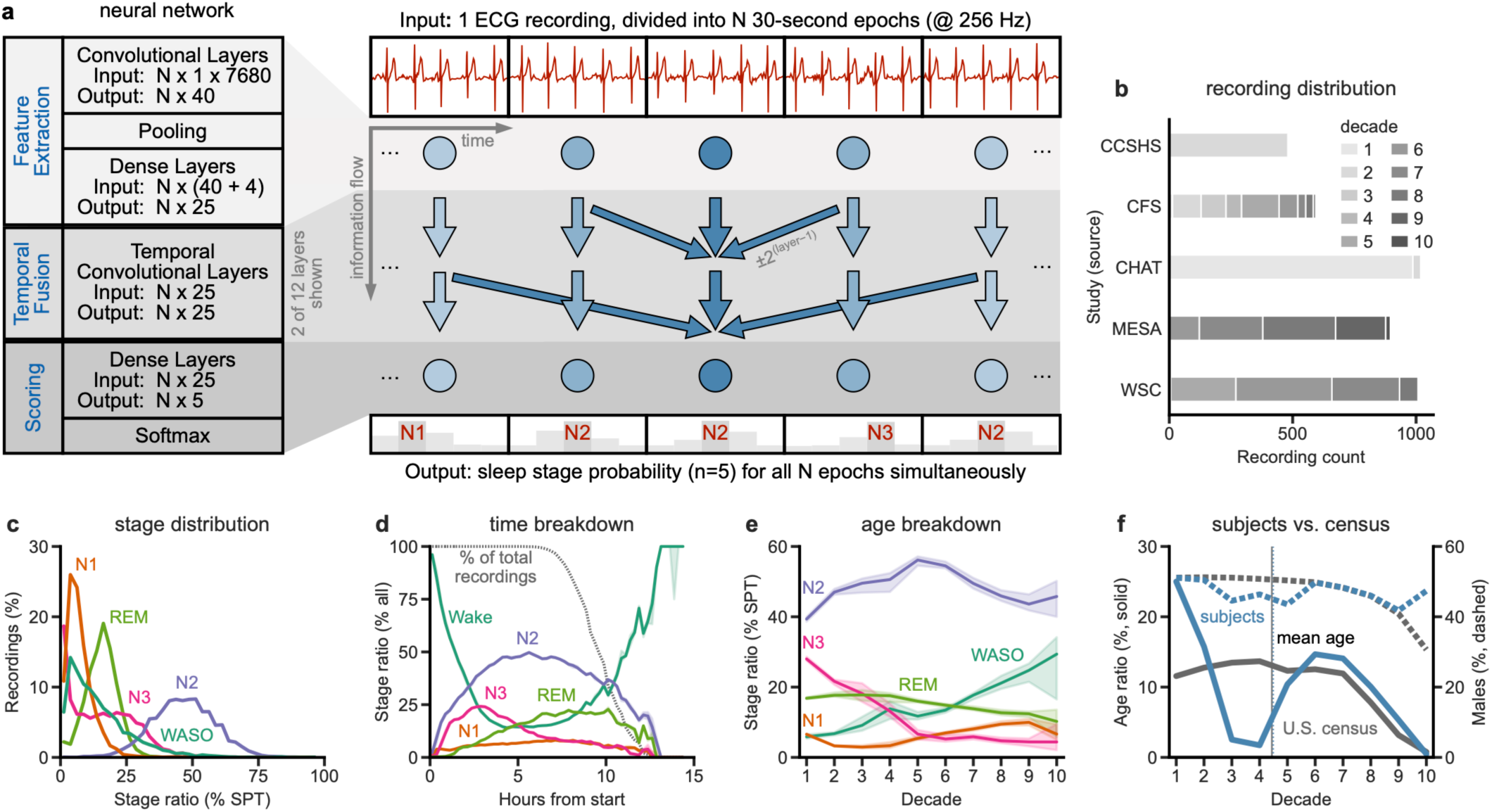
| Network architecture and input. (a) The network consists of three groups of layers. It scores all epochs (N) in a recording simultaneously, with ellipses representing duplication of network structure across the recording. The four other inputs (not shown) are age, sex, wall time, and relative epoch position within the recording. Most arrows are hidden for clarity, and ECG and epoch stage scores are for illustrative purposes only. (b) The 4,000 recordings came from five studies, with the distribution of the subject’s age in decades shown. (c) There is a wide distribution of stage ratios as a percentage of sleep period time (SPT, i.e., the period between and including the first and last epoch of sleep). Wake after sleep onset (WASO) is any wake (arousals) during SPT. (d) As expected from previous studies, recordings show time-dependent changes in the relative proportion of the various sleep stages. E.g., N3 is more likely to occur at the beginning, and REM is more likely to occur near the end. e) The data also show expected age-dependent changes in sleep. In particular, the ratio of time spent in stage N3 declines with age, whereas arousals (WASO) increase. (f) We aimed to select subjects (blue lines) to match the U.S. census statistics (gray lines) by age (solid lines) and sex (dashed lines). The lack of subjects in decades 3-5 is a limitation of the available datasets (decade 1=age 0-9yr.), with subjects added to other decades to achieve the same mean age as the census data.

The network (Fig. 1a) takes as input a complete recording of normalized ECG data, the subject’s sex and current age, as well as the wall time (i.e., clock time) and the relative position of each recording epoch (described in detail in Methods). The network then computes 25 features for each 30-second epoch, fuses those features across time, and finally outputs the probability of each of the five sleep stages for each epoch simultaneously. The network classifies the epoch as the stage with the highest probability.

### Comparison with expert human-scored PSG and EEG-less models

To assess the claim that the model achieved expert-level human performance, we need to compare its performance against how well human scorers agree with each other. Specifically, we compared the model’s performance with published summary statistics from the large SIESTA dataset, for which we have inter-rater reliability^12,11^. It is reasonable to posit that where the model’s median performance overlaps or exceeds the human inter-rater reliability, it performs at or above expert human level, respectively. Using this criterion, the model (n = 500, testing set on ECG) performs on par with expert human scorers (n = 72, SIESTA PSG) overall (Fig. 2a), and for stages Wake and N3 (Fig. 2c). The boxplot notches represent the 95% CIs for the medians (see Methods).

**Figure 2.**
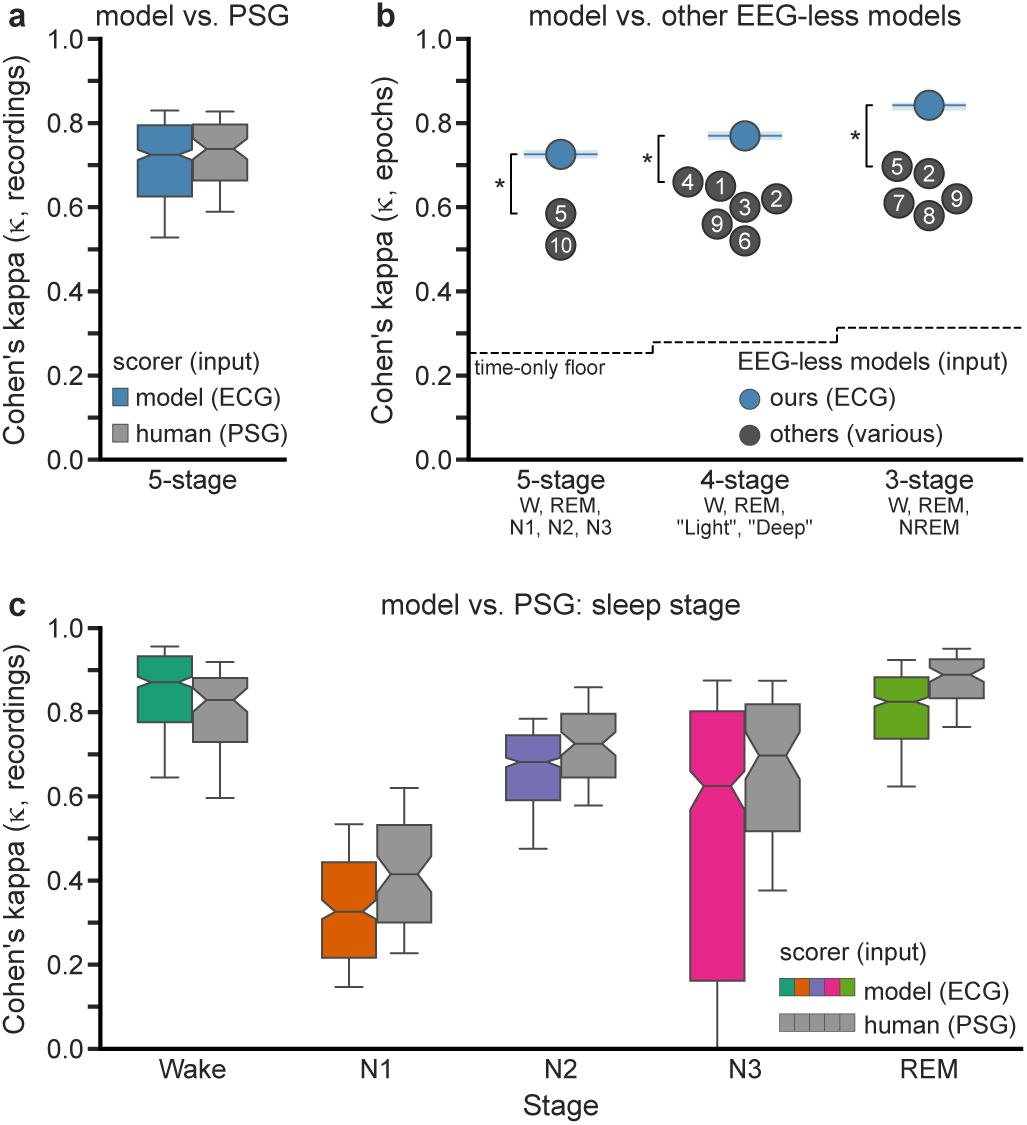
| Performance comparisons against human-scored PSG and EEG-less models. (a) Five-stage Cohen’s kappa (κ) for our model based on single-lead ECG (blue) and human-scored PSG (gray). Overlapping notches indicate that the network performs on par with expert human-scored PSG. Model kappas were based on the hold-out testing set of n=500 recordings from distinct subjects, and human kappas were calculated on an n=72 dataset scored with R&K^12^. (b) Our model (blue dot and horizontal line) performs significantly better than other EEG-less models (black dots with numbers), regardless of stage granularity—even while using fewer inputs (* percentile bootstrap p < 0.0002). The bootstrapped 95% CIs were smaller than the dot diameters (shown as shading, see table for values). The other dots represent the kappa values of the recent best (κ≥0.5) EEG-less algorithms and are listed in Extended Data Table 2, with models sorted by year. The sources are: 1) Radha^17^ 2) Wulterkens^18^ 3) Fonseca^19^ 4) Sridhar^20^ 5) Sun^13^, 6) Beattie^21^, 7) Yoon^22^, 8) Domingues^23^, 9) Willemen^24^, 10) Sady^25^. Note that our algorithm uses ECG as the only input, while many other models also use respiration, actigraphy, or both. A “time-only floor” shows the performance of the current model structure when only using two time variables (wall clock time and relative epoch position), i.e., no ECG or demographic data. All kappa values are shown in Extended Data Table 1. (c) Additional stage-specific results for comparison against human-scored PSG on the same data and methodology as panel a. Whiskers are at P10 and P90. Notch and bootstrap calculations are in Methods.

Additionally, we compared our algorithm against other recent best (κ ≥ 0.5) EEG-less algorithms and devices—including the state-of-the-art (Fig. 2b). Because most published algorithms use more coarse stage granularity, we also trained and evaluated the final model for four-stage (Wake/“Light”/ “Deep”/REM) and three-stage (Wake/NREM/REM) scoring by combining the appropriate stages (see Methods). The algorithm performs significantly better (p < 0.0002) than published EEG-less algorithms that have reported kappa values for five-stage scoring (1=0.726 versus 0.585^13^—evaluated on all epochs). Furthermore, the algorithm performs better regardless of the granularity of stages (e.g., five, four, or three-stage scoring) or the number of additional inputs used (e.g., actigraphy, respiration, HRV, etc.; Extended Data Table 2).

We also evaluated the model on additional recordings to test if there were unique attributes about the recordings selected or if study-specific learning had occurred—which would hamper generalizability. The first collection consists of recordings from the original five studies that met the quality criteria (see Methods) but which we had not randomly selected to be part of the 4,000 recordings (Extended Data Figure 2a). It bears stressing that we did not add these recordings to our 500-recording testing set because this would skew the age and sex distributions from the target census distribution. These results show no significant difference between those recordings that were selected or unselected. The second collection comes from a study, MROS, which we did not use for the training, validation, or testing sets. Their performance was equivalent to the age-matched males (decades ≥ 7) in the testing set (Extended Data Figure 2b). This result indicates that if the model had learned any study-specific features, they were unnecessary for adequate performance.

Another way to assess how well the network agrees with human-scored stages is to compute the contingency table. The row-normalized contingency table (similar to a confusion matrix) showed excellent agreement, with an overall agreement of 80.0% with human-provided scores (Fig. 3a). The one exception was with N1, which is often scored as one of the stages that naturally precede or follow it in time, namely Wake and N2—matching what has been reported elsewhere with human scorers of PSG data^12^. Furthermore, we found the expected pattern for all stages; when there was disagreement, it was scored as the stage that typically preceded or followed it.

**Figure 3.**
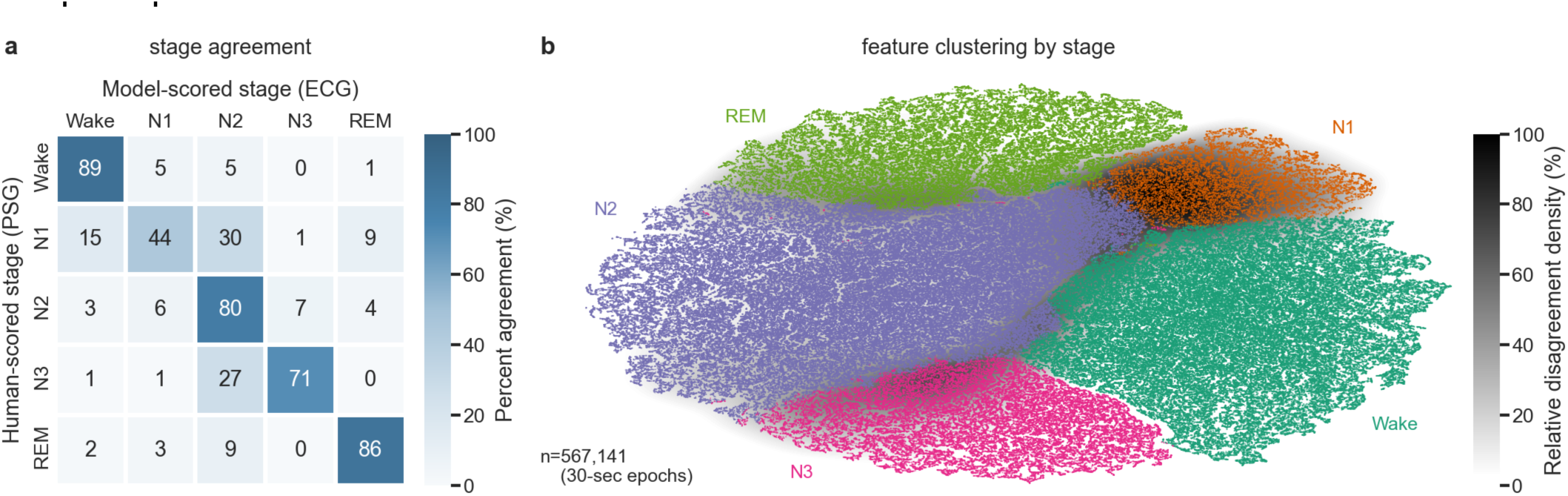
| Normalized contingency table and t-SNE of all epochs. The row-normalized contingency table of all n=567,141 epochs shows the high degree of agreement between human- and model-scored stages. When disagreement exists, the scorer usually labels the stage as the one that typically precedes or follows it during the night; e.g., N1 is scored as Wake or N2, N3 is scored as N2, and REM is scored as N2. The lowest agreement occurs with N1, consistent with human scorers of PSG data. Overall agreement is 80.0%. (b) t-SNE plot taken from the 25 outputs of the penultimate layer of the network. Dots represent the epochs where the model agrees with the human-scored stage. Due to the sparsity of the disagreements, a kernel density estimate is used to show where the disagreements occur (black = highest disagreement density). The greatest disagreement density occurs when scoring N1 epochs. Disagreement typically exists along each of the cluster borders except Wake-N3.

We confirmed these findings by investigating the network’s feature space, specifically by creating a t-SNE plot (Fig. 3b) of the 25 features for each epoch that came from the penultimate layer of the network (i.e., before the final classification occurred). The t-SNE calculations reduce the feature space to two dimensions and place similar epochs in the 25-D space closer together in the 2-D space. We plotted the epochs where the human and model agree as dots, colored by stage. Compared with the agreements, the disagreements are less frequent (in n = 453,866 epochs, the model agreed with humans, while in n = 113,275 epochs, it did not). Therefore, we used a kernel density estimate to show the density of the disagreements. Similar to the normalized contingency table (Fig. 3a), the greatest disagreement occurs when scoring N1 epochs, especially near the boundary between N1 and N2. The disagreements also occur along boundaries between adjacent stages in a typical recording.

In addition to assessing the overall performance of the network (i.e., Cohen’s kappa) to compare the model with human performance, we also examined sleep stage transitions. The overall, Wake, and N1 median transition rates are equivalent for the model and human (Fig. 4a). The overall transition rate also aligns with results reported by others^14^. Although the boxplots for the various stages largely overlap, N3 transitions show the greatest difference. Furthermore, the transition matrices for the model and human scores (Fig. 4b,c) are nearly identical, with a bootstrapped Pearson’s r of 0.998.

**Figure 4.**
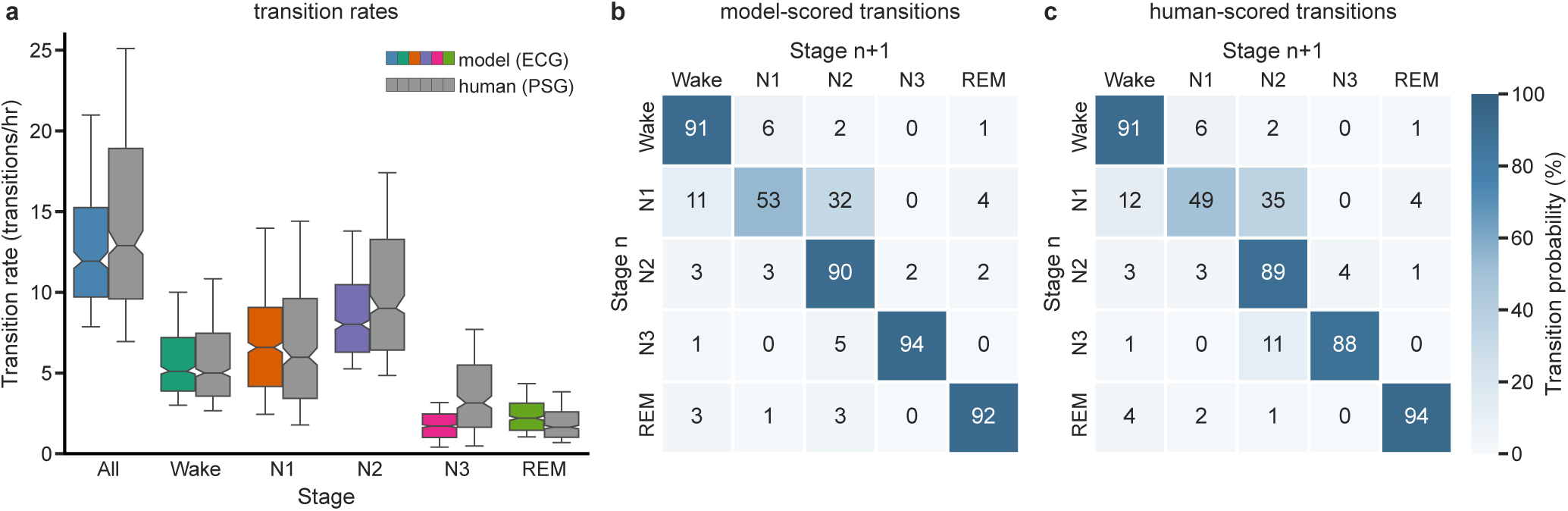
| Stage transitions. (a) Transition rates for all five sleep stages for the n=500 testing set for our ECG-based model (various colors) and the human-scored PSG (gray) as an additional measure of human-model agreement. There is no difference in the median transition rates for All, Wake, and N1, significantly fewer transitions for the model for stages N2 and N3, and significantly more transitions for stage REM. (b) The transition matrix of the model’s classifications for all n=567,141 epochs. (c) The transition matrix of the human’s classifications (same n). The Pearson’s r of the two matrices (panels b and c) is 0.998, with percentile bootstrapped 95% CI [0.997, 0.999]. Overall, the next epoch is likely the same stage as the current one. Consistent with the literature, the probability that N1 will transition into a different stage (namely N2 or Wake) is higher than for other stages. Whiskers at P10 and P90.

Next, we assessed the network’s performance on both the original testing set and specific modifications to it to help better understand the results in Fig. 2 (e.g., to determine why any differences exist) and to determine under what conditions the network is best suited and when and why it might perform worse than expected.

The initial series of results were on the original testing set. First, the network’s performance is largely unaffected by age (Fig. 5a) except for N3, which decreases beginning in the fifth decade. This result is expected and reported elsewhere^11^, likely because with age, sleep becomes shorter and more fragmented^15^. Consistent with this, we further found that performance deteriorated as a function of sleep stage ratio. Specifically, when there were very few or too many (i.e., nearly all) epochs of a given stage, the model performed less well (Fig. 5b). Similar to the finding on stage ratios, the amount of pre-sleep wake (i.e., time awake between when the recording started and the subject fell asleep) was important (Fig. 5e, the distribution of pre-sleep wake is shown in Extended Data Figure 1c), as performance deteriorated when there were fewer than 30 minutes of pre-sleep wake.

**Figure 5.**
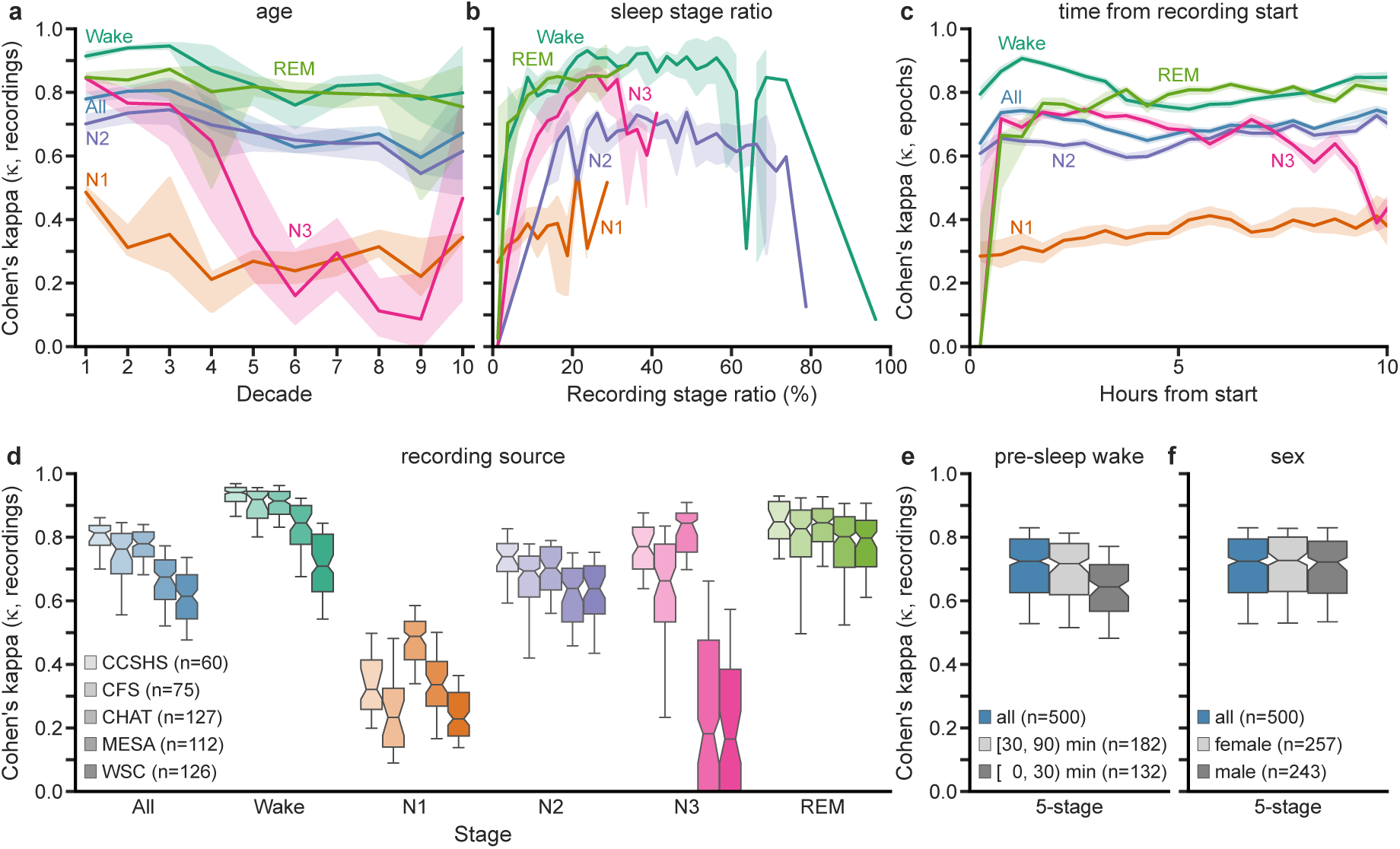
| Performance on the testing set. (a) When stratified by age (decade 1=age 0-9yr.), the overall kappa is only slightly affected by age, while N1 and N3 are the most affected, suggesting the broad applicability of the model. (b) Stratified by the recording’s stage ratio, namely the proportion of recorded time spent in a given stage, all stages perform significantly less well when their stage ratio is minuscule—or nearly everything. In these situations, model results should be interpreted with caution. (c) When stratified by time (30-min windows across all recordings), although there is some variation in kappas, the stage-specific kappas fall in a narrow range—matching the overall performance boxplots (Fig. 2a,c). However, the lower model performance is because of a lack of diverse observations in those periods (e.g., REM is rare at the beginning of the night, whereas N3 becomes less frequent after eight hours Fig. 1c). (d) Stratified by dataset, stage-specific kappas vary little, except N3—which is significantly lower for MESA and WSC (which consist primarily of older subjects, Fig. 1b). (e) Stratified by the amount of pre-sleep wake, kappa is significantly lower for recordings with less than 30 minutes of pre-sleep wake; prospective users of the model should plan accordingly. Most recordings (n=314) contain less than 90 minutes. (f) Stratified by sex, kappas show no differences, suggesting equal applicability of the model to both sexes. Shaded areas represent the 95% CI. Whiskers at P10 and P90.

On the other hand, although the likelihood of a particular stage varies across the night, it has little effect on performance (Fig. 5c), except on the classification of N3 and REM right at the start of the night and N3 after about 9 hours. Around the same time, the network begins to score most of the human-scored N3 epochs as N2 (Extended Data Figure 1b). Performance did depend on which study it came from (Fig. 5d), as illustrated by significantly worse N3 performance for recordings from MESA and WSC (which also contained most of the older subjects). Finally, the sex of the subject did not affect the performance (Fig. 5f, Extended Data Figure 1d).

### Robustness to missing data, noise, and other perturbations

Next, we investigated the limits of the network and its suitability by modifying the testing set and examining how it affected performance. Because we cannot generate synthetic ECG data corresponding to a particular sleep stage, we worked with additions to or deletions of existing data. We stress that we performed all these experiments on the final model and never trained the network on these manipulations (e.g., we did not add noise during training to make the model more robust).

First, we examined the effect of trimming the recordings, either from the beginning (Fig. 6a) or from the end (Fig. 6d). When trimming from the beginning, the performance for all stages except N1 immediately trends downward, albeit to varying extents. When trimming from the end, however, the performance was nearly unaffected until after we removed many hours of data. The result underscores the greater importance of the initial period of the recording for classifying sleep stages (echoing pre-sleep wake effects, Fig. 5c). To assess the network’s ability to handle noise (e.g., environmental), we separately added four different sources of noise: white Gaussian noise, and three from the MIT-BIH noise stress test^16^ namely, baseline wander, electrode movement, and movement artifact. For each of these noise types, we added the noise in increasing amounts, up to 100% of the signal’s power (i.e., SNR = 0 dB).

**Figure 6.**
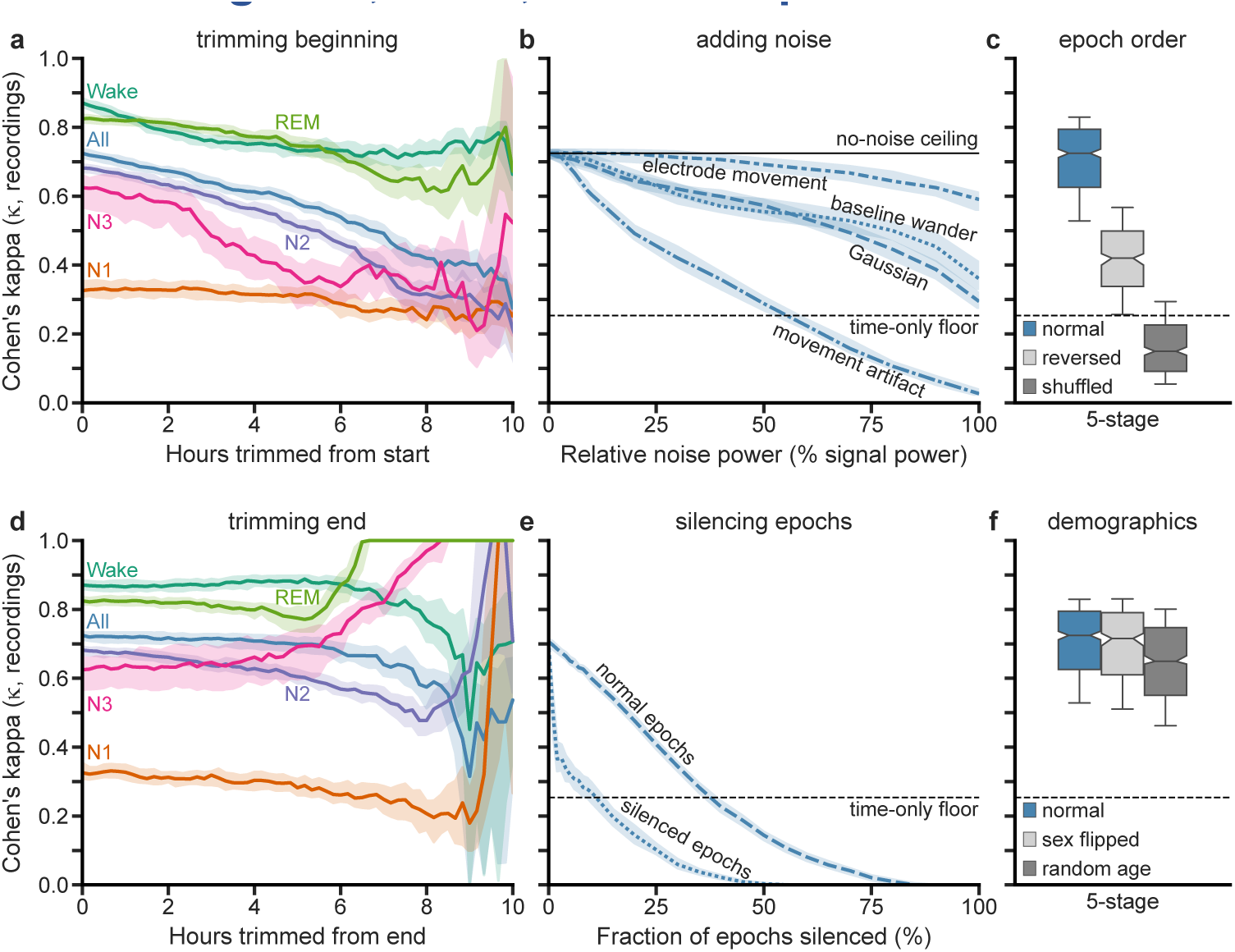
| Performance with noise or modifications to the testing set. (a) When trimming recordings from the start, there is a gradual downward trend in kappa for all stages except N1, indicating the importance of the early periods. (b) Likewise, performance decreases with relative noise power (noise power divided by signal power) for four different noise sources (electrode movement, baseline wander, Gaussian, and movement artifact). Shown are a “no-noise ceiling” (zero noise added, black solid line) and a “time-only floor” (see Fig. 2b, black dashed line, which continues into panel c). The model exhibits remarkable robustness for all noise sources tested except movement artifact, as the performance exceeds the floor, even at 100% relative noise. (c) When the epoch order of each recording is reversed or shuffled, the performance is significantly worse, indicating that the model strongly relies on temporal order. (d) When trimming the recordings from the end, the performance is less affected than when trimmed from the start (panel a), indicating it will perform well even if the recording stopped prematurely. (e) Performance as a function of the fraction of all epochs silenced (replaced with zeros) quickly trends downward, as the model uses context for both intact and silenced epochs, suggesting that the model will suffer if electrodes are disconnected for extended periods. (f) There is no change in performance when the subject’s sex is flipped, suggesting some insensitivity to sex. In contrast, performance is significantly lower when a random age is assigned. Shaded areas represent the 95% CI. Whiskers at P10 and P90.

Furthermore, we scaled the noise relative to each epoch’s signal power to compensate for variations across the recording. These experiments (Fig. 6b) demonstrate that substantial noise levels are required to affect the performance substantially. The exception is movement artifact noise, which causes a steeper roll-off. Moreover, with increasing movement artifact noise, the network increasingly scores epochs as Wake (Extended Data Figure 1a). Next, because we designed the network to use context, it is helpful to evaluate what happens when the context is drastically modified by reversing or shuffling the epoch order (Fig. 6c). Reversing the epoch order significantly affects the performance negatively, and even more so when epoch order is shuffled. We also evaluated how losing entire epochs, possibly due to loose leads, affects the model’s performance. For scale, an average of four minutes per recording (and a maximum of 3.6 hr.) of the ECG data used in this study was lost to loose leads (Methods). To simulate this, we silenced (i.e., set the signal to a value of 0) individual epochs selected randomly from the recording (Fig. 6e) and analyzed the kappas of the intact and silenced epochs separately. Model performance decreases for both, indicating that the network uses the ECG input from surrounding epochs in determining the sleep stage for a given epoch (Fig. 6c)—and yet is robust enough to lose up to 35% and still exceed the “time-only” floor. Finally, we evaluated the usefulness of the two demographic inputs by either flipping the sex for all subjects or assigning an age with uniform probability from 5 to 90 years old (Fig. 6f). We see there is a significant effect for modifying age but not sex.

Lastly, we silenced portions of the input for each epoch to determine what portions of an individual epoch the network uses in constructing its features. The results show that the network is more affected by losing the data from the epoch’s center (dotted line) than from its ends (dashed line), and the network can lose about 5% from each end (10% total) before the performance begins to degrade (Extended Data Figure 1e).

### Real-time operation

Note that all the results presented above scored the entire night of sleep simultaneously, which means information from past and future epochs contributed to classifying the sleep stage. Similar to the computations performed when we silenced random epochs (Fig. 6e), computations of the normalized relative importance (Methods) of epochs showed that the network is using contextual information before and after to score that epoch (Fig. 7b). Of note are the “blips” at powers of two that reflect the underlying structure of the network (Fig. 1b). We wondered how well a (slightly modified) network could score sleep in real time. Therefore, we altered the structure of the network such that it could only use information from the current and past epochs (Fig. 7a). From examining the real-time network, we found that although the performance is slightly lower, there is no statistical difference overall between the real-time network and either the base network or human-scored PSG (Fig. 7c).

**Figure 7.**
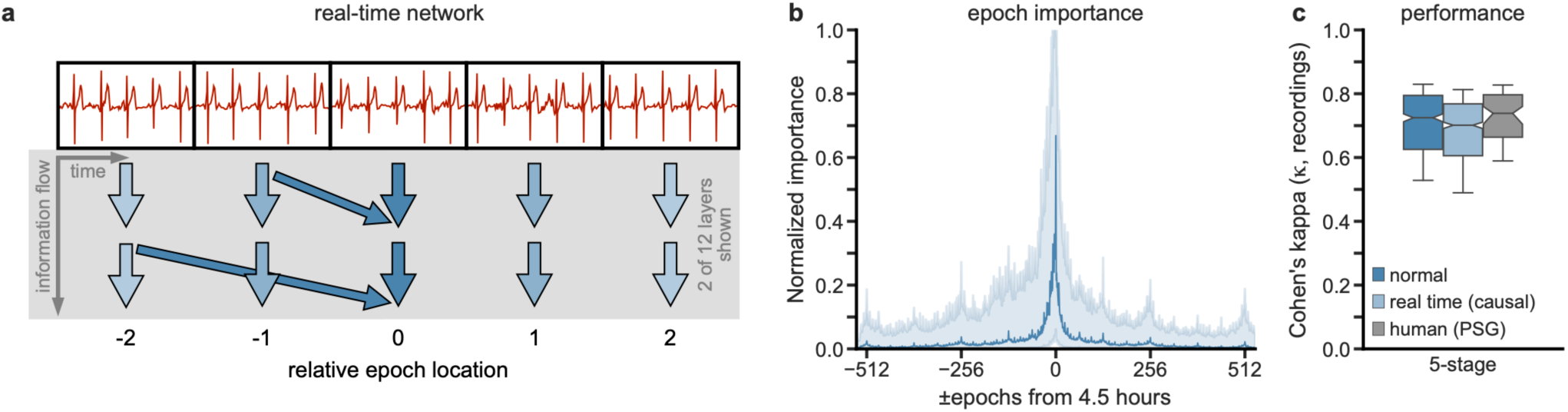
| Real-time network and epoch importance. (a) The “Temporal Fusion” section of the network (Fig. 1a) was modified to make the network score in real-time (i.e., causally). Note how no arrows point anti-causally. Most arrows are hidden for clarity. (b) Using integrated gradients (Methods) to determine the relative importance of the epochs used to score at epoch 540 (4.5 hours from the start) shows that the baseline network (Fig. 1a) makes use of past and future epochs, with noticeable “blips” at powers of two distance from current. The median importance of n=500 recordings is shown, and shaded areas represent the 95% CI. (c) The overall performance of the real-time (causal) is trending lower (but not significantly) than that of the original (non-causal) network and also than human-scored PSG (the notches overlap). This result is a desirable property for potential real-time applications for our approach. Whiskers at P10 and P90.

## Discussion

To the best of our knowledge, our study represents the first successful implementation of five-stage sleep staging comparable to expert-scored PSG without the aid of EEG (or the host of other sensors used by PSG). Similar EEG-less studies have shown correlations between sleep stages defined by PSG and data obtained from other non-EEG sensors. The advantages of replacing human scorers with automated algorithms are significantly reduced labor costs and increased inter-rater reliability. In addition to these benefits, utilizing ECG instead of PSG introduces further advantages, such as a more user-friendly setup, a more robust signal, and broader accessibility to the scientific community as well as citizen scientists. Prior to our work, the suboptimal performance of EEG-less methods in five-stage classification had suggested that EEG would always be necessary for achieving clinically relevant sleep staging. However, our findings demonstrate that ECG-based automated sleep staging can achieve comparable performance to PSG-based human sleep staging, thereby challenging the notion that EEG is indispensable.

### On par with human-scored PSG

Regarding the performance of our model, it is noteworthy that it achieves expert-human level agreement overall (Fig. 2a) and for Wake and N3 (Fig. 2c). Moreover, the consistency between the network’s classifications and the human-scored stages is evident when analyzing the row-normalized contingency table (Fig. 3a), stage transition rates (Fig. 4a), and stage transition matrix (Fig. 4b,c). These findings strongly support the argument that our network scores sleep stages similarly and on par with human-scored PSG. Like human scorers, the network also incorporates contextual information (Fig. 6c,e). Finally, in a slightly modified form, it can perform real-time scoring (Fig. 7c)—also at a level equivalent to offline human-scored PSG.

However, we observed that the median performance of the model for REM sleep, while very high (1 = 0.825 on all recordings), was nonetheless statistically worse than human-scored PSG (Fig. 2c) since the 95% CIs do not overlap the human-scored intervals. One possible explanation for this discrepancy is the absence of inputs from eye movement (i.e., EOG) and muscle activity (i.e., EMG), as even PSG, relying solely on EEG, struggles to classify REM sleep reliably.

Although the model’s performance in classifying N3 is equivalent to human-scored PSG, further discussion is warranted. Stratifying the results by their source dataset reveals that the network performs significantly worse on the MESA and WSC datasets (Fig. 5d), primarily due to decreased kappas for Wake and N3. The drop in N3 kappa could be partially attributed to the worse N3 performance with age (Fig. 5a), observed for human-scored PSG as well^11^.

Additionally, the fact that MESA was studying atherosclerosis—which by definition negatively impacts cardiac physiology—and that both studies included older subjects with lower-than-expected N3 amounts for their age range^10^ may have contributed to worse-than-expected N3 kappa. Furthermore, the performance across time (Fig. 5c) demonstrates that, for most stages, the model’s performance is relatively insensitive to time during the recording (except at the ends)—except perhaps N3 and REM. Notably, the network begins predominantly scoring epochs classified as N3 by human scorers as N2 after approximately 9 hours (Extended Data Figure 1b), indicating a potential qualitative difference in N3 sleep occurring at the end versus earlier in the night.

Our model significantly outperforms other EEG-less models, irrespective of five-, four-, or three-stage scoring (Fig. 2b). While the current literature on EEG-less methods (including recent commercial sleep-tracking devices) is more extensive than what is referenced here, papers were excluded for one or more of the following: i) their results did not list kappa or did not include sufficient data to calculate it; ii) they only considered two-stage (Wake/Sleep) scoring, which has limited clinical or research utility; or iii) their evaluation set was non-independent of their training set (see Supplemental Discussion for details).

Finally, our model demonstrates robustness to added noise and data corruption, which is advantageous in variable and harsher environments outside the clinic. Specifically, the model was able to perform very well to common and harsh noise sources—even with relative noise powers exceeding 50% (Fig. 6b). Moreover, the results indicate that the model performs best when the subject has the relatively non-intrusive ECG strap attached at least 30 minutes before falling asleep (Fig. 5e). However, the performance is not contingent upon knowing when sleep began and ended.

### Autonomic nervous system

ECG serves as a valuable non-invasive tool for observing the activation of the autonomic nervous system. The branches of which, namely the sympathetic and the parasympathetic systems, are known to have characteristic effects on heartbeats and heartbeat rates. Although the interactions between the autonomic and central nervous systems are bidirectional, traditionally, it has been assumed that autonomic activity usually follows central activity and that autonomic is subordinate to the central system. However, extensive research has shown that autonomic activity sometimes precedes central activity, such as in decision-making and attention^26^. Additionally, novel statistical measures like transfer entropy have revealed that during sleep, more information flows from the autonomic to the central nervous system than in the reverse direction. Furthermore, specific activations of the autonomic nervous system during sleep reliably lead to distinct activity in the central nervous system, both in terms of cortical location and frequency content of EEG^27^. The fact that our ECG-based neural network performs comparably to PSG in determining sleep stages aligns with findings of a healthy interaction between the autonomic and central nervous systems and the idea that sleep is important for the brain and body.

### Limitations

There were numerous differences in equipment and aims between the studies we used. These differences were partially due to the lower importance of ECG during PSG and included electrode placement, sampling rates, and quantization. In addition to harmonizing these differences where possible (which brings about its own issues), we had to discard a sizable percentage of recordings because of poor data quality (Methods). Finally, because we resampled all data to 256 Hz for our model, we could not determine the effect of the sampling rate on performance.

Because it is still unknown what exactly characterizes the ECG of a given sleep stage, it is not yet possible to generate data synthetically. This limited our ability to explore the robustness of the network to only adding noise or modifying the recordings (e.g., silencing epochs), as opposed to synthesizing atypical heartbeats or rhythms. Relatedly, along with the lack of ground truth labels, we could not explore purposeful misclassifications by modifying an epoch to “look” more like another stage—except for movement artifact noise (Fig. 6b), which had the effect of eventually making all epochs look like wake (Extended Data Figure 1a).

As mentioned, the data we used came from studies that scored their data using either R&K or AASM. Moreover, although there are slight differences in the criteria for the similarly named stages, it was necessary to harmonize them. The refinements in the scoring criteria lead to some differences—even when scorers use both methodologies for the same recording^28^. While the total sleep time (i.e., Wake vs. sleep) and REM scoring will be nearly identical, others have found that for the non-REM stages (i.e., N1/N2/N3), the distributions will slightly change—a decrease in N2 (∼5%), which is distributed nearly-equally to N1 and N3. Because we used a single class label for each similarly named stage, the network likely had to make compromises where the two methodologies would disagree.

Finally, and most foundationally, this work is based on R&K and AASM scoring rules and the orthodoxy that sleep can be cleanly segmented into discrete stages that are, at least, self-consistent. However, many alternative methods used to score sleep and critiques of these rules exist^29^, as well as data that supports a more heterogenous—both across and within a stage— understanding of sleep stages^30^.

### Cardiosomnography

The effectiveness of ECG in sleep medicine for detecting sleep disorders, such as apnea, has been demonstrated by numerous studies^27,31,32^. Building upon these findings, we propose that cardiosomnography—a sleep study conducted with electrocardiography only—can complement and supplement PSG in sleep medicine. Compared to PSG, cardiosomnography offers advantages in terms of cost-effectiveness and simplicity.

The adoption of cardiosomnography has the potential to facilitate more accessible sleep medicine, thereby enabling further research, interventions, and applications. For instance, it has been found that in Alzheimer’s disease, a feedback loop can arise where the progression of the disease can lead to increasingly disturbed sleep^33^. In turn, chronic poor sleep can contribute to the accumulation of β-amyloid^34^. Continuous and cost-effective sleep monitoring and disease progression tracking for at-risk individuals could guide treatments and potentially improve outcomes.

Additionally, cardiosomnography could be a valuable tool for interventions. Research has indicated that playing pink noise only during N3 increases the amplitude of slow oscillation waves and the proportion of N3 during the night^4,35^. Bringing this intervention out of the clinic with real-time ECG-based sleep staging could be helpful for those with mild cognitive impairments, who typically exhibit reduced amounts of N3^36^. Furthermore, for consumer applications, it has been found that entering and quickly exiting N1 can enhance creativity^37^. With a less cumbersome method of monitoring and alerting users during N1, developers could make this intervention widely available.

In summary, cardiosomnography holds great potential to benefit researchers, physicians, and entrepreneurs by more easily providing objective measures of sleep for interventions and sleep medicine as a whole. Moreover, it opens up possibilities for as-yet-unknown applications and could expand the scope of sleep-related research, interventions, and healthcare.

## Methods

### Datasets

In order to build a model that is useful for a wide range of human ages and a variety of health conditions, we used data from five large datasets from the National Sleep Research Resource^38^: Cleveland Children’s Sleep and Health Study (CCSHS)^39^, Cleveland Family Study (CFS)^40^, Childhood Adenotonsillectomy Trial (CHAT)^41^, Multi-Ethnic Study of Atherosclerosis (MESA)^42^, and Wisconsin Sleep Cohort (WSC)^43^. These datasets consist of PSG recordings scored using either R&K (CCSHS, CFS, and WSC) or AASM (CHAT and MESA).

In addition to the wide range of ages (5 to 90 years), these datasets provided diversity in sex, race, ethnicity, and medical conditions. We did not exclude any recordings based on subject characteristics (i.e., demographics, health, medications, etc.).

### ECG data

Even though the recordings included data from many biophysical inputs, we used only ECG lead I (the limb lead across the heart) from each recording for our model’s input. Some studies provided ECG lead I as a single channel of ECG data (often labeled “ECG” or “EKG”). And for other studies, we calculated ECG lead I by subtracting the signal provided by the right (RA, often written as “ECG1”) and left (LA, often written as “ECG2”) limb electrodes.

### Pre-processing

There was a considerable variation in data quality in the ECG recordings because ECG is rarely the channel of interest for PSG (e.g., loose leads, poor connections, different sampling rates, and environmental noise). We had to process and evaluate all 8,611 recordings provided by the five datasets to determine which recordings we could use to train and evaluate the network. To that end, all recordings went through an automated pre-processing algorithm described below. It bears mention that we took the recordings as-is and did not trim wake periods before they fell asleep (mean sleep latency = 1.3±1 hr.—one s.d., see Extended Data Figure 1c) or after they woke up.

If the ECG length was not a multiple of 30 seconds, we trimmed it down to the next nearest 30-sec epoch length.

We silenced sections where leads had come loose. If the ECG lead came from two electrodes, we silenced both signals whenever either electrode came loose.

If we used two electrodes, we subtracted one from the other to obtain ECG lead I.

High-pass filter (0.5 Hz) the data to attenuate baseline wander but maintain longer features, such as T waves.

Remove 60 Hz line noise with a notch filter.

Remove any additional constant-frequency noises using notch filters.

Resample to a single common frequency (256 Hz).

Normalize using a robust z-score.

### Detecting heartbeats

Once we had pre-processed all the data, the next step was to roughly identify most heartbeats in each recording. This step involved finding heartbeats in each recording based on archetypical heartbeat templates. Then, we generated a recording-specific template for each recording from the first pass of detected heartbeats. Finally, we performed a second pass across the recording to add or remove heartbeats that matched the recording-specific template.

Once we had identified the putative heartbeats for each recording, we could calculate the recording-specific normalization factor, which brings all recordings to the same scale. To do this, we calculated the maximum absolute value for every heartbeat previously identified. Then we took the 90% percentile of all maximum values as the maximum threshold. We then doubled that threshold to obtain the normalization factor. We then divided the ECG obtained after the pre-processing by this normalization factor. The result will be that all, or nearly all, heartbeats will fall within the range of a value of ±0.5. Finally, to eliminate extreme values (which will make training more difficult), anywhere the ECG exceeds ±1, we clipped it to ±1.

### Acceptable recording criteria

We wanted to ensure that all the data used to train and evaluate the model were of good quality. Therefore, we set selection criteria based on the ECG only—we used no criteria based on the stage scores (e.g., time spent awake, etc.).

At least 5 hours of data, but no more than 15 hours.

Contain a lead I ECG channel or two electrode channels that we could use to derive it.

A sampling rate of at least 100 Hz.

There were three or fewer constant-frequency noises (including 60 Hz).

At least 85% of the epochs must contain possibly useable data (defined as having more than eight unique values).

At least 85% of the epochs must contain at least one template-matching heartbeat.

At least 85% of the epochs must contain median absolute deviation (MAD) values ≥ -3 s.d. of the robust z-scored MAD values for all epochs.

The normalization factor must be ≤ 250.

After dividing by the normalization factor, ≤ 5% of the data can be extreme values (those outside ±1).

### Recording selection and set building

At this point, there were 5,718 recordings remaining (of the original 8,611) which met the above selection criteria. Unfortunately, it was obvious that there would not be sufficient recordings from subjects of the 3^rd^, 4^th^, 5^th^, and 10^th^ decades such that we would be able to match the U.S. age and sex distribution as per the current U.S. census estimates. Therefore, we instead over-sample subjects from the remaining decades with the following goals: the mean age should match that of the U.S. census, and the subjects from the 6-9^th^ decades should be over-sampled by the same number. Furthermore, we also desired to match the sex distributions for each decade, as provided by the census data.

As a compromise on training time versus volume of data, we chose to select a total of 4,000 recordings that we would use for the training, validation, and testing sets. We used random sampling to select the 4,000 recordings from the 5,718 available—with the age and sex distributions specified above. Because there are more recordings than unique subjects, we put additional criteria in the random selection process: the testing set must contain 500 unique subjects. Furthermore, although we allowed the validation and training set to contain more than one recording from the same subject, we required each set to have a unique set of subjects that were not shared with any other set. Finally, because this random selection process could draw from the original datasets unequally, the last step was to shuffle recordings between the sets to achieve similar ratios of the source datasets across sets. It bears stressing that we did not add the 1,718 recordings that the algorithm had not explicitly selected to our 500-recording testing set because this would skew the age and sex distributions from the census distribution.

The final distribution of the 4,000 recordings included 4,597,343 epochs, or 38,311 hours, of data (duration = 9.6±1.4 hr.). See Extended Data Table 3 for the set-specific counts), and Fig. 1 for visual representations of the recording distributions.

### Scores and weights

The sleep score annotations were provided in various file formats and, as mentioned above, using either R&K or AASM scoring criteria. Although slight differences exist in the similarly named stages, we harmonized the annotations with the following two adjustments to the annotated scores. When a dataset scored with R&K provided separate S3 and S4 stages, we combined them into a single stage: slow wave sleep (SWS/N3). Second, if the human scorer had annotated an epoch as “unscored”, “movement”, or anything except the five stages of interest, then we made the following changes: we changed the score to Wake, and we set the weight of the epoch to zero. In total, 1.2% of the epochs fell into this bucket. Setting the weight to zero prevented the network from being penalized for not correctly predicting the stage. In addition, because the human annotator had not scored these epochs as one of the five stages, we excluded them from all results.

We also adjusted the epoch-specific weights when the data quality of the epoch was poor. If we had silenced a portion of an epoch due to loose leads, we set the weight to one minus the proportion removed. Therefore, if we had silenced an entire epoch due to a loose lead, it received a weight of zero. Finally, if an epoch contained eight or fewer unique values (i.e., distinct voltage readings), we considered it devoid of signal and assigned a weight of zero. In all, 0.9% of the epochs had no ECG data. In contrast to the unscored epochs above, we still included these in the kappa calculations, although they were devoid of data.

### Performance metric: Cohen’s kappa

To compare the algorithm’s performance on scoring sleep stages, we need to use an established metric. When classifying sleep stages, there is some subjectivity and therefore no “ground truth” label for any given epoch; even identically trained sleep scorers will have some disagreement^12,11^. Therefore, the most appropriate statistic is the inter-rater reliability, or the degree of agreement between two observers. We used Cohen’s kappa (1), the most commonly reported inter-rater reliability statistic in sleep research.

Cohen’s kappa measures the agreement between two raters that cannot be attributed to chance alone. The statistic uses the probability of observed agreement (p_o_) and the probability of chance agreement (p_e_): 1 = (𝑝_e_ − 𝑝_*e*_)⁄(1 − 𝑝_*e*_), where a value of 1 = 0 means no agreement above chance level, while 1 = 1 means perfect agreement—where both raters match on all labels. The minimum value depends on the marginal distributions and is between zero and -1.

Although it was designed for binary classification, to compute the individual class-specific kappas in a multiclass task such as sleep staging, the contingency table is often split into separate class-versus-others tables for each class^44^. For instance, to compute kappa for N3, one can use N3 as the first class and combine all other classes (Wake+N1+N2+REM) into an aggregate class.

It should be noted that if there are two or more classes, and both raters score everything as just one of the classes, the naïve formulation will produce an undefined value. However, in this case, both raters perfectly agree (i.e., 1 = 1); therefore, we set kappa to 1.

Depending on the field, two different methods of computing Cohen’s kappa exist. Therefore, we must do the same to compare our results with others. The first method, often used by human-scored PSG literature, computes kappa for each recording individually and calculates summary statistics on those kappa values. The second method, favored by machine learning literature, aggregates one contingency table for all the epochs from all recordings and then computes kappa on that aggregated contingency table. We found that when the number of epochs and recordings are both large—such as in the present study—the median kappa of all recordings is nearly identical to the kappa of all epochs. However, for all results, we specify which method we used. We have reported numerical values for kappa, as there is no agreed-upon standard for quantitative labels.

### A new kappa-correlated loss function

A differentiable loss (or objective) function is necessary for a neural network to learn, i.e., to calculate the gradients used to adjust the weights and biases. Our criterion for the loss function was the highest overall kappa with the narrowest possible range of individual stage-specific kappas. In other words, every stage-specific kappa be as high as possible, instead of the typical outcome where the classifier largely ignores the minority classes.

For a classification task such as sleep staging, cross-entropy loss (aka log-likelihood) is the standard^45^. However, the first issue we had with cross-entropy is that it assumes that the classes are nearly equal in size. When class size imbalances exist, such as with sleep stages, cross-entropy often disregards the minority classes. The issue is significant for N1, which often constitutes less than 5% of the night but is still an essential marker of the wake-sleep transition. Various techniques are employed to overcome this issue^46^, including under- and over-sampling the classes. However, a data-level solution was not possible here because our network simultaneously scores an entire night of sleep, making it impossible to balance the proportions of the classes artificially. Another solution often used is weighting each class’s importance. Extensive evaluations demonstrated that this was also inadequate; although the accuracy slightly increased, it had a negligible effect on the kappa of N1.

The second related issue with standard classification loss functions is that they assume accuracy is the intended performance metric. However, accuracy is only loosely correlated with kappa. Although 1 = 1 when accuracy is 100%, there is no one-to-one mapping below this, and the exact value will depend on the relative proportions of the classes. For instance, when accuracy is 50%, kappa could be almost any value between -1 and 1. That is to say, for the same accuracy, kappa could be wildly different.

Due to the limitations of existing loss functions, we developed a new function. The new loss function is one minus the geometric mean of the scaled class-specific kappas. As expected, our loss function (termed the class kappa mean) is (negatively) correlated with the overall kappa, which is the weighted arithmetic mean of the class-specific kappas. An advantage to using the geometric mean is that it is more invariant to the relative proportion of the classes, i.e., the function is less likely to ignore the minority class. Since kappa can technically range from -1 to 1, the formula re-scales the kappas to stay within the range [0, 1] to prevent issues that may arise with a kappa less than or equal to zero. For the formula (Equation 1), *c* is the current class, and *n* is the number of classes (for five-stage scoring, *n* = 5).

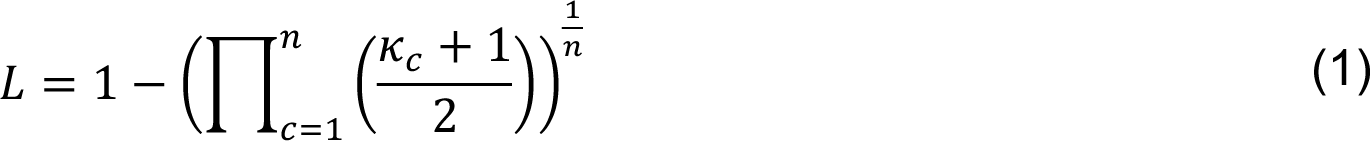

We calculated the kappas for the loss function using the contingency table generated from the output of the softmax layer (described below in Neural network)—which returns an output that resembles a confidence value for each class. The training algorithm builds the contingency table from the softmax outputs, which enables the back-propagation algorithm (which uses the loss function) to guide the network to be more confident in the correct predictions while minimizing the likelihood and confidence of incorrect predictions. Results comparing this new loss function against others are in Extended Data Table 4. The new loss function was critical for training the neural network described below.

### Neural network

The neural network takes the input data (i.e., ECG, age, sex, time) and produces sleep stage probabilities for all epochs. Conceptually, we divided the network’s layers into three groups (see Fig. 1 for a simplified network representation). The first group, the feature extraction layers, extracts relevant features from the input data for each epoch. The second group, the temporal fusion layers, combines the features temporally across epochs. The third group, the classification layers, uses the fused features to predict the probability for each stage of each epoch.

First, the feature extraction layers take the input to produce a set of features. The inputs include the cleaned, but otherwise untransformed, ECG (described above), the subject’s sex (Boolean value) and age at the time of recording (normalized to 1 = 100 yr.), epoch’s relative location within the recording (−1 = beginning, 1 = end), and wall time (where 0 = midnight and ±1 = ±24 hr.). The wall time is included because circadian rhythms influence stage proportions^47^. The ECG data passes through several convolutional and pooling layers to extract 40 features. Then the network combines those ECG features with the subject’s age and sex, as well as the time variables. Finally, several dense linear layers reduce the output to 25 features for each epoch.

Next, the temporal fusion layers take the 25 features for each epoch and merge them across time. The configuration is based on the temporal convolution network^48^, with a modification to merge information before and after the current epoch. The first layer merges the features from the current epoch and epochs before and after it (epoch±1). Then the second layer merges the already-merged features from the current epoch and the epochs located two positions before and after it (epoch±2). This motif continues upward for 12 layers as epoch±2(layer−1), such that any epoch in the top layer could combine information from epoch±34 hr.—significantly longer than any recording used.

Finally, the classification layers take the 25 features from the final temporal fusion layer through several layers of dense linear units. The final softmax layer transforms the predictions into an output that resembles confidence for each of the five sleep stages.

### Training and evaluation

Designing a neural network involves a hyperparameter tuning process (e.g., determining the optimal number of layers, node types, non-linearities, etc.). During the hyperparameter search, we only trained the model on the training set (n = 3,000) and only evaluated it on the validation set (n = 500). We used a separate validation set because cross-validation (i.e., partitioning the data such that every sample is eventually evaluated) is inappropriate during hyperparameter turning—as it leads to information leaking into the model. A more detailed description of the history of our hyperparameter search and the problem with cross-validation during hyperparameter tuning is in the Supplement Methods.

In addition to standard techniques to improve regularization, we used automatic per-recording weighting during training, which downweighted recordings with notably lower kappas. We did this to prevent the network from misusing parameters to improve classification on just a handful of outlier recordings. To improve the network’s robustness to inverted leads (i.e., the ECG leads had been attached to the wrong terminals by the technician—a problem that we identified had occasionally happened), we had the data loader invert the ECG of every recording 50% of the time during training—but not during evaluation.

The network was trained using a variant of the Adam optimizer with stable weight decay, AdamS^49^. The learning rate automatically decreased whenever the performance plateaued on the evaluation set. Additionally, our training algorithm reduced the learning rate by half when performance plateaued for 50 epochs on the evaluation set. The initial learning rate was 1×10^-3^, with a minimum of 1×10^-6^. We trained the model using a batch size of 10 (i.e., the number of recordings grouped together) for a maximum of 1,000 training epochs (i.e., iterations over the entire dataset). We stopped the hyperparameter search and selected the final model when we decided that additional changes were unlikely to significantly improve the results on the validation set.

After we selected the final model, we retrained it on the joint training and validation set (n = 3,500) and evaluated it on the hold-out testing set (n = 500)—which we had never used before that point.

## Statistics

Where possible, we did not assume a normal distribution for any of the results. Instead, we extensively used nonparametric bootstrapping—a method that makes no assumptions about normality. For all bootstrapped results, we always used a sample of the same size as the original (e.g., n=500 for the testing set) with 10,001 iterations. Moreover, we use the median, a more robust measure of central tendency, as the estimator. All 95% CIs are estimated using percentile bootstrapping of the median. Finally, whenever line charts show shaded regions, the line is the median, and the shaded regions are the 95% CIs.

## Analyses with the five-stage model

Most of the analyses and results we computed used a single five-stage model that we trained once (described above) and then evaluated on the original testing set and any modified versions of that set described below.

### Comparison with other expert-scored PSG

Unfortunately, only a handful of studies have conducted more than a cursory investigation of the inter-rater reliability of human PSG scorers. The SIESTA dataset was collected with this investigation in mind, and it is the most extensive study to calculate inter-rater reliability^10,11,12,50^. However, we only have access to the summary statistics provided.

Therefore, to compare the performance, it made the most sense to compare the median kappas for each stage of the model on the testing set against results reported on SIESTA. Furthermore, where the model’s median performance overlaps—or exceeds—the human inter-rater reliability, the model performs at or above expert human level. The comparison study comprises 72 recordings from 56 healthy controls and 16 subjects with different sleep disorders (mean age: 57.7±18.7, 34 females)^12^.

To determine the 95% CIs of the medians for the SIESTA data, we used the standard formula: ±1.573𝐼𝑄𝑅⁄√𝑛9^51^. This formula assumes their data is normally distributed—which we were unable to confirm. For the model’s testing set, we used bootstrapping to determine the 95% CIs of the median. Finally, we can conclude with 95% confidence that the true population medians differ where the CIs of a given pair of medians do not overlap. In other words, we cannot reject the hypothesis that there are no differences in the medians where the CIs overlap.

### Comparison with EEG-less models

To compare our results on the five-stage model with the other EEG-less algorithms, we calculated Cohen’s kappa on a bootstrapped aggregate contingency table. If the next-best performing model is below the lowest bootstrapped result, we can only conclude that the p-value is below two times the inverse of the bootstrap iteration count.

However, since most models reported in the literature do not evaluate five-stage scoring, we aggregated the stages from the five-stage model’s results (e.g., for four-stage scoring, setting epochs of either N1 or N2 to “Light” sleep) and performed the same bootstrap operation described above to compare the four- and three-stage performance.

### Comparison with the human-provided scores

To compare the human-score stages with the model-scored stages, we used a row-normalized contingency table to show where there is the most substantial agreement and disagreement. To look at the clustering of the features that the network builds to score sleep, we used a t-distributed stochastic neighbor embedding (t-SNE) to reduce the 25 features down to two—which is more easily plotted. Doing so allowed us to visualize clusters in the embedding space.

Furthermore, we can see where these disagreements lie in this 2-D representation by transforming the epochs that the human and model disagree on. In addition to the overall agreement (i.e., Cohen’s kappa) between the human and model, we also investigated the transitions between stages—specifically, the transition rates and probabilities. Moreover, we compared the human and model transition matrices by bootstrapping and using Pearson’s r to calculate a correlation between them.

### Robustness analyses

To analyze the robustness of the network, we conducted several experiments (given the limitation that we could not generate new data): we trimmed recordings, added noise, modified each recording’s epoch order, silenced whole (and portions of) epochs, and change the original demographics. We trimmed the recordings by removing epochs from the beginning or end. For the noise experiments, we individually added four different sources of noise: white Gaussian noise, as well as three noises from the MIT-BIH noise stress test^16^ (i.e., baseline wander, electrode movement, and movement artifact).

Several of these experiments allow us to investigate how much contextual information matters. Silencing (or setting to zero) whole epochs was the first experiment. We randomly selected epochs in each recording and silenced their ECG input. Another experiment to investigate the contextual information was either flipping the epoch order of each recording or shuffling the order. Each epoch would still have the ECG progressing through time in the correct direction, but adjacent epochs would now be out of order. Finally, using the method of integrated gradients^52^, we can investigate how important each epoch in a recording is for scoring a given epoch.

Finally, we evaluated the remaining two inputs: sex and age. By either flipping the sex or assigning a random age, we can determine how important those variables are to the performance of the network. Related to the integrated gradient analysis of the epochs, silencing portions of all epochs allowed us to investigate what portion of the epoch the network was using. Either we silenced the epoch from both ends or the center.

## Analyses with other models

As mentioned, most of the results presented come from just the single five-stage model described above. However, we trained an additional eleven models; we used two models to gain additional network insights, and we used nine models to compare and assess the loss function we presented. We stress that the single five-stage model above stands alone, and the additional models do not form an ensemble.

### Determine a time-only floor

If one were to remove the ECG input entirely, it would be reasonable to suspect that the model would perform no better than chance (1 = 0). However, the fact that sleep stages occur in cycles and have an expected progression across the night indicates that it might be possible to use temporal information alone to achieve better-than-chance agreement. We were unaware of literature evaluating this idea and decided to determine what kappa is with only the two time variables as valid input (i.e., wall time, and relative epoch position). To minimize changes in the network structure, instead of removing the unnecessary convolution and pooling layers, we replaced the ECG, age, and sex data with random data that was sampled each time a recording was loaded. We trained (one) time-only five-stage model.

### Real-time operation

A scorer typically scores sleep after the night or recording is over. Therefore, the scorer has access to the entire night to assist them in classifying each epoch. However, having real-time (i.e., causal) scoring would be necessary for some of the interventions and applications we believe would benefit from cardiosomnography. To that end, we trained (one) five-stage real-time model that we had modified so that for any given epoch, it could not use information from the epochs that followed it (i.e., the future). This modification included removing (by replacing it with random data) the relative epoch position input, as this would’ve revealed how much time was left to the network.

### Compare the new loss function

Although we developed the new loss function during the hyperparameter search, it would be helpful for interested users to know how well other commonly used loss functions could perform on the same five-stage model. To that end, we trained the same five-stage model (four) times with different loss functions. Furthermore, because of the tradeoffs that all loss functions exhibit, we wanted to determine what would happen if the same five-stage model only had to score a single stage (e.g., only Wake vs sleep). Therefore, we also trained (five) five-stage models with our loss function, but only to optimize one of the five stages.

### Evaluation of additional hold-out recordings

As mentioned, there were an additional 1,718 recordings from the original five studies that met the acceptable recording criteria but which we did not add to the training, validation, or testing sets. We did this out of a desire to limit training time and to match the U.S. census distribution. However, we did evaluate these recordings separately to determine if there was anything unique about the recordings we had randomly selected.

Furthermore, we evaluated recordings from a dataset we did not use in the training phase, namely the MrOS Sleep Study (MROS)^53^. We analyzed these recordings to determine if the model was learning and using any study- or site-specific features from the five primary studies. The study provided 3,933 recordings, of which 3,193 met the quality criteria.

## Data availability

All human input data for this study came from publicly available datasets at the National Sleep Research Resource (https://sleepdata.org/).

## Code availability

All code and model weights will be made available the date of publication.

## Author contributions

A.M.J. and B.R.S. conceived the study. A.M.J. designed and trained the deep network, analyzed the data, produced the results, and wrote the manuscript. B.R.S. regularly assisted, including with discussing results and revising the manuscript. L.I. provided critical feedback on the results and text. All authors discussed the results and contributed to the final manuscript.

## Data Availability

All human input data for this study came from publicly available datasets at the National Sleep Research Resource (https://sleepdata.org/). All code and model weights will be made available the date of peer-reviewed publication.

## Acknowledgments

The authors acknowledge the use of the Opuntia, Sabine, and Carya clusters and the support from the Research Computing Data Core at the University of Houston to carry out the research presented here.

The Cleveland Children’s Sleep and Health Study (CCSHS) was supported by grants from the National Institutes of Health (RO1HL60957, K23 HL04426, RO1 NR02707, M01 Rrmpd0380-39). The National Sleep Research Resource was supported by the National Heart, Lung, and Blood Institute (R24 HL114473, 75N92019R002).

The Cleveland Family Study (CFS) was supported by grants from the National Institutes of Health (HL46380, M01 RR00080-39, T32-HL07567, RO1-46380). The National Sleep Research Resource was supported by the National Heart, Lung, and Blood Institute (R24 HL114473, 75N92019R002).

The Childhood Adenotonsillectomy Trial (CHAT) was supported by the National Institutes of Health (HL083075, HL083129, UL1-RR-024134, UL1 RR024989). The National Sleep Research Resource was supported by the National Heart, Lung, and Blood Institute (R24 HL114473, 75N92019R002).

The Multi-Ethnic Study of Atherosclerosis (MESA) Sleep Ancillary study was funded by NIH-NHLBI Association of Sleep Disorders with Cardiovascular Health Across Ethnic Groups (RO1 HL098433). MESA is supported by NHLBI funded contracts HHSN268201500003I, N01-HC-95159, N01-HC-95160, N01-HC-95161, N01-HC-95162, N01-HC-95163, N01-HC-95164, N01-HC-95165, N01-HC-95166, N01-HC-95167, N01-HC-95168 and N01-HC-95169 from the National Heart, Lung, and Blood Institute, and by cooperative agreements UL1-TR-000040, UL1-TR-001079, and UL1-TR-001420 funded by NCATS. The National Sleep Research Resource was supported by the National Heart, Lung, and Blood Institute (R24 HL114473, 75N92019R002).

The National Heart, Lung, and Blood Institute provided funding for the ancillary MrOS Sleep Study (MROS), “Outcomes of Sleep Disorders in Older Men,” under the following grant numbers: R01 HL071194, R01 HL070848, R01 HL070847, R01 HL070842, R01 HL070841, R01 HL070837, R01 HL070838, and R01 HL070839. The National Sleep Research Resource was supported by the National Heart, Lung, and Blood Institute (R24 HL114473, 75N92019R002).

This Wisconsin Sleep Cohort Study (WSC) was supported by the U.S. National Institutes of Health, National Heart, Lung, and Blood Institute (R01HL62252), National Institute on Aging (R01AG036838, R01AG058680), and the National Center for Research Resources (1UL1RR025011). The National Sleep Research Resource was supported by the U.S. National Institutes of Health, National Heart Lung and Blood Institute (R24 HL114473, 75N92019R002).

## Extended Data

**Extended Data Figure 1.**
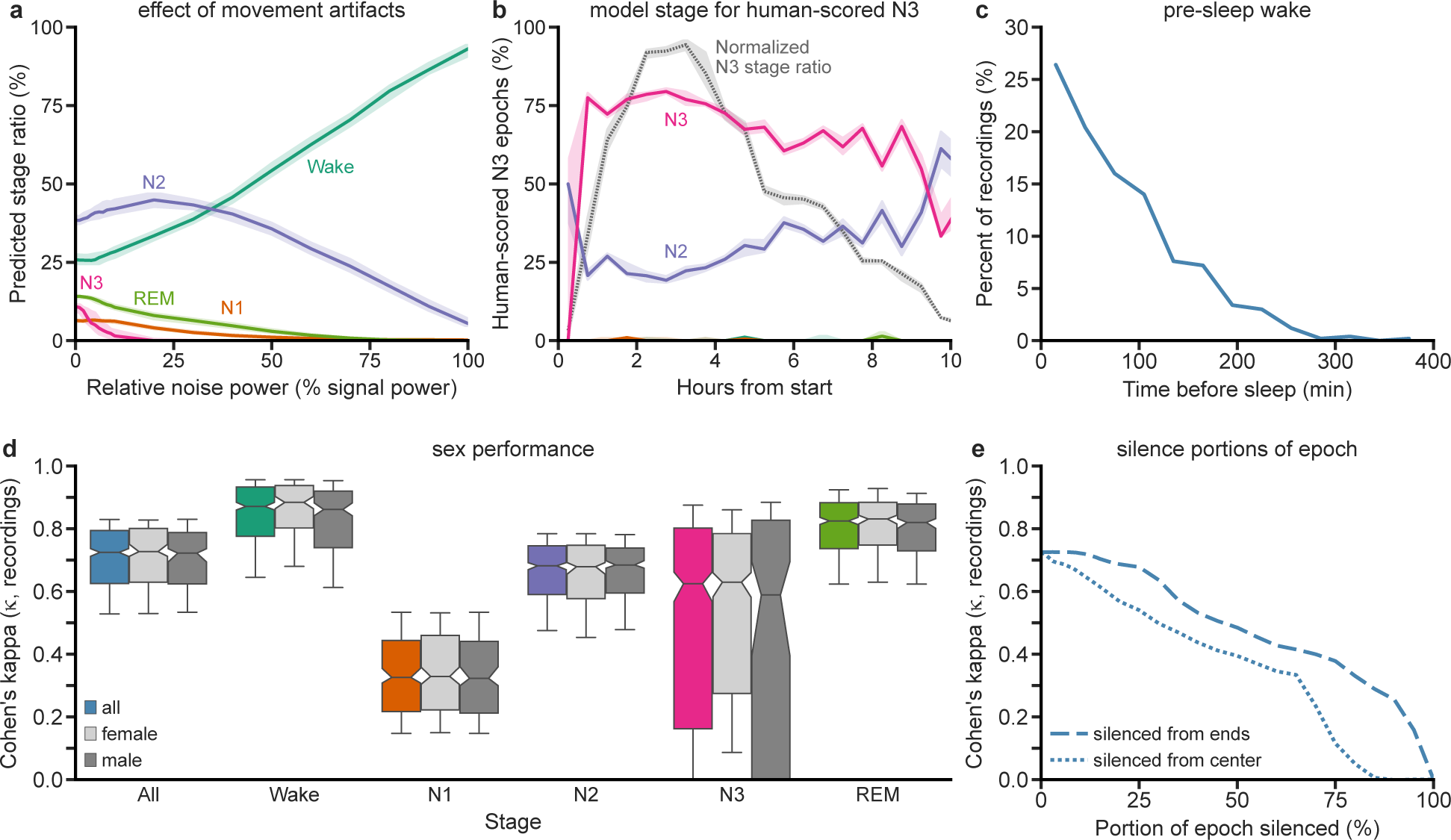
| Additional plots. (a) When adding movement artifact noise (see Fig. 6b), with increasing noise power, the network scores more epochs as Wake. (b) Looking only at human-scored N3 epochs, the model predominantly scores them as N3 until around 9 hours in and then as N2. The gray line is the normalized likelihood of N3 epochs (i.e., where in the night N3 was most often being scored by the human, Fig. 1d). (c) A majority (n=314) of recordings contain less than 90 minutes of pre-sleep wake. (d) When stratifying by sex, results are essentially the same for each stage, with noticeable but insignificant sex differences for Wake and N3. (e) When silencing increasing portions of each epoch, starting from both ends of each epoch, performance is degraded with increasing silence. Performance is worse when silencing from the center than from the ends. Shaded areas represent the 95% CI. Whiskers at P10 and P90.

**Extended Data Figure 2.**
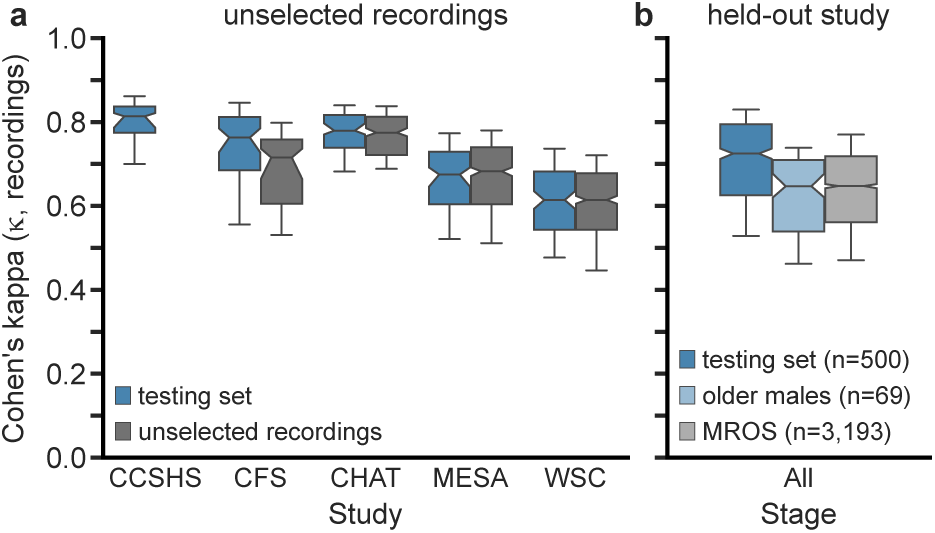
| Unselected recordings and held-out study. (a) For five of the studies from which recordings were drawn, four were not used in their entirety (see Methods). The results of those unselected recordings show no differences from their counterparts in the testing set. CFS (n=105), CHAT (n=270), MESA (n=498), WSC (n=845). (b) One study, MROS (n=3,193), was not used during the training phase, so we could evaluate it to determine if study-specific learning had occurred (i.e., learning features specific to the study apparatus or pipeline). The performance aligns with the aged-matched males that constituted the study’s demographics (decades ≥ 7) from the testing set (light blue). Whiskers at P10 and P90.

**Extended Data Table 1.** | Cohen’s kappas on testing set.

| Kappa calculation method | Cohen's kappa ($\kappa$) | | | | | |
| --- | --- | --- | --- | --- | --- | --- |
|  | All | Wake | N1 | N2 | N3 | REM |
| Median kappa of all recordings (n=500) | 0.725 | 0.871 | 0.326 | 0.682 | 0.625 | 0.825 |
| Kappa of all epochs (n=567,141) | 0.726 | 0.862 | 0.373 | 0.671 | 0.703 | 0.805 |

Depending on the method used to aggregate the results, there are slight differences in values reported. Both are shown here to enable direct comparisons with a wider variety of literature.

**Extended Data Table 2.**
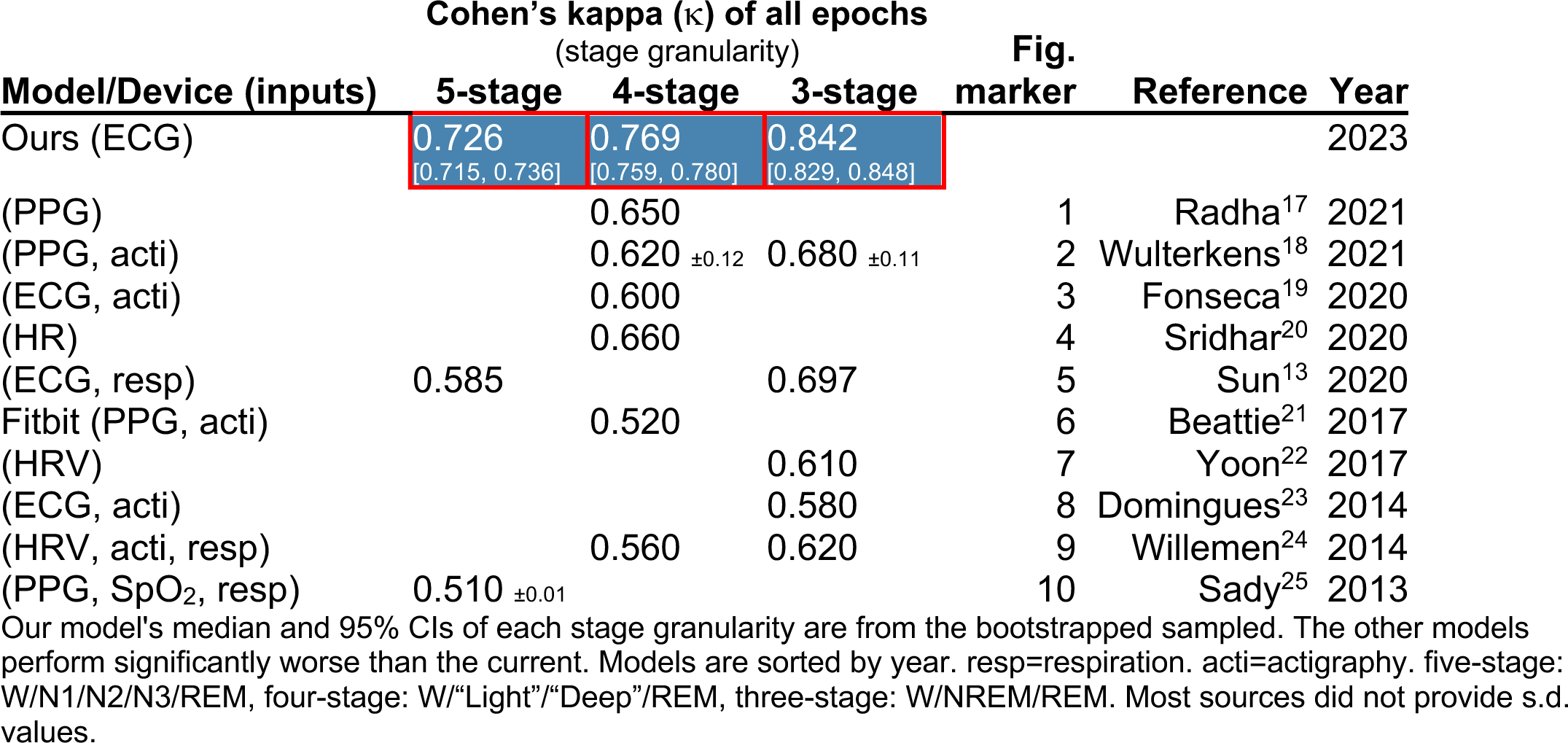
| Comparison with other EEG-less algorithms.

**Extended Data Table 3.** | Epoch counts for each set Epoch counts for each stage.

| Set | Epoch counts for each stage |  |  |  |  |  | Total |
| --- | --- | --- | --- | --- | --- | --- | --- |
|  | Wake | N1 | N2 | N3 | REM | Unscored |  |
| Training | 980,338 | 209,550 | 1,315,130 | 457,009 | 447,811 | 41,387 | 3,451,225 |
| Validation | 165,513 | 34,830 | 218,264 | 74,034 | 71,904 | 7,223 | 571,768 |
| Testing | 165,300 | 34,601 | 220,642 | 73,566 | 73,032 | 7,209 | 574,350 |
| Total | 1,311,151 | 278,981 | 1,754,036 | 604,609 | 592,747 | 55,819 | 4,597,343 |

For the 4,000 recordings selected, above are the epoch counts of each stage and those that were unscored.

**Extended Data Table 4.** | Loss function comparisons.

| Loss function | Cohen's kappa ($\kappa$) of all epochs | | | | | |
| --- | --- | --- | --- | --- | --- | --- |
|  | All | Wake | N1 | N2 | N3 | REM |
| Class kappa mean (ours) | 0.726 | 0.862 | 0.373 | 0.671 | 0.703 | 0.805 |
| Cross-entropy | 0.734 | 0.867 | 0.274 | 0.682 | 0.699 | 0.805 |
| Cross-entropy (weighted) | 0.669 | 0.845 | 0.332 | 0.583 | 0.677 | 0.786 |
| Focal<sup>54</sup> | 0.732 | 0.862 | 0.297 | 0.679 | 0.703 | 0.801 |
| Cohen's kappa (overall) | 0.720 | 0.854 | 0.000 | 0.669 | 0.697 | 0.795 |
| Ratio of ours to best | 99% | 99% | 100% | 98% | 100% | 100% |

Although some loss functions achieve slightly better overall performance, they do so with much lower N1 performance. The highest performance for each column is highlighted in gray. The weights provided for weighted cross-entropy were the inverse of the stage proportions in the n=3,000 training set (i.e., [0.214, 1.000, 0.159, 0.459, 0.468]).

**Extended Data Table 5.** | Five-stage versus one-stage comparisons.

| Loss function | Cohen's kappa ($\kappa$) of all epochs | | | | |
| --- | --- | --- | --- | --- | --- |
|  | Wake | N1 | N2 | N3 | REM |
| Complete 5-stage | 0.862 | 0.373 | 0.671 | 0.703 | 0.805 |
| Best 1-stage (each) | 0.862 | 0.353 | 0.671 | 0.692 | 0.800 |
| Ratio to 1-stage | 100% | 106% | 100% | 102% | 101% |

When the loss function uses all five stages simultaneously (i.e., the geometric mean), it performs equal to or better than any stage-specific kappa that might be optimized individually.

