## Supplementary Information for "Electrocardiogram sleep staging on par with expert polysomnography"

### Supplementary Methods

#### Hyperparameter search

Not only was the final network organically grown and modified during the hyperparameter search (e.g., adding layers, changing the number of features, etc.), but we also changed the inputs to the network. Before we settled on the basic structure of using a fully feed-forward network, we had also tried traditional machine learning techniques (support vector machines, naïve Bayes, etc.) and, later, a recurrent neural network. These earlier attempts used many hand-crafted inputs that we assumed would be necessary based on our extensive signals processing experience and a review of similar attempts. The current iteration of the network initially included the spectrum of the ECG for each epoch and the autocorrelation of seven, 4-second windows for each epoch (to approximate HRV). However, later we found that the performance improved slightly if we removed the spectrum and autocorrelation inputs—leaving ECG as the only biophysical input.

### Supplementary Discussion

#### Loss function comparison

Since we developed our loss function during the hyperparameter search, the results back then on each of the loss functions would make little sense when compared with the results presented here. Therefore, we re-ran the training using the final model with the three best-performing loss functions and the overall Cohen's kappa. It is important to remember that the loss function probably influenced the network's evolution, and therefore the final network might be less performant on any other loss function. However, the reported results mirror the relative values we saw during development. Overall, during the hyperparameter search, we examined several dozen ways of combining and weighting various loss functions to little avail. Most loss functions either completely ignored N1 (i.e., $\kappa = 0$) or could not bring N1's performance up to what we eventually found was achievable with our loss function.

The results in Extended Data Table 4 demonstrate that although unweighted cross-entropy and focal loss can achieve slightly better performance in terms of the overall kappa (+1%, both) or the kappas for some stages, the N1 performance is significantly worse (-62 and 59%, respectively). Given that the new loss function had significantly better N1 performance versus the marginal decrease in overall performance, we decided it was a worthwhile tradeoff.

#### Exclusion of some EEG-less studies

The final issue with some EEG-less studies mentioned in the Discussion, contamination of the evaluation set, describes a serious methodological problem that comes in two forms and is surprisingly common. The first form of this issue is using so-called "subject-specific" classifiers. The researchers trained and evaluated these models on data from the same recordings, whereby an individual epoch was included in either the training or evaluation set. The problem is that the data will be nearly identical between adjacent or nearby epochs. Therefore, the evaluation data is highly similar to the training data. The second form of the issue is using a single evaluation set; there should be two evaluation sets, a validation set, and a hold-out testing set, which the model should never see until the final evaluation. During the development of any model, hyperparameter tuning is necessary to achieve the best-performing model. To improve the model and converge on the best hyperparameters, researchers use a validation set that is different from the training set. However, they also sometimes perform this step using cross-validation. The problem is that the hyperparameter tuning process “leaks” information from the validation set into the model (i.e., the researchers make model choices based on the performance on the validation set). Furthermore, when evaluating the model for generalizability, i.e., the performance on unseen data, if the same validation set is used for testing (or the same cross-validation population), the researcher is unwittingly evaluating the performance against data already seen. Reviewing the literature requires carefully reading the methods to notice these issues. It is often only obvious when the results specifically mention an “external” or “unseen” data evaluation, where the performance is usually significantly worse than their top-line numbers.

### Future directions

The ability of a neural network to score sleep stages using a single lead of ECG on par with experienced human scorers using data from a dozen or more electrodes raises several questions. The most salient question is what specifically in the input data is the network using to such a pronounced effect. As mentioned, other EEG-less algorithms have been mining downstream measures of ECG, such as HRV, with limited success compared to PSG performance. Moreover, in an earlier iteration of our network, we used additional inputs, including a surrogate for HRV—with no improvement in performance. We would like to investigate what the network is using.

Finally, it is worth highlighting that we only took one preemptive measure to improve the network’s robustness while training. While spot-checking the input ECG data, we noticed from the waveform appearance that some electrodes had been connected backward (i.e., the polarity was reversed). Instead of manually verifying all recordings, we inverted the ECG during training with a 50% probability—as mentioned in the Methods. This operation undoubtedly made learning more difficult, forcing the network to develop a polarity-insensitive feature extraction. This preemptive technique could be used in future iterations to improve the network's robustness further. Specifically, Gaussian, or other forms of noise, could be added, or portions of epochs or even entire epochs could be removed. We emphasize that there will likely be tradeoffs between incorporating these measures and the training time and final performance.
